# Social connections at work and mental health during the first wave of the COVID-19 pandemic: Evidence from employees in Germany

**DOI:** 10.1101/2022.03.04.22270966

**Authors:** Jonas Breetzke, Eva-Maria Wild

## Abstract

Empirical evidence on the social and psychological impact of the COVID-19 pandemic in the workplace and the resulting consequences for the mental health of employees is still sparse. As a result, research on this subject is urgently needed to develop appropriate countermeasures. This study builds on Person-Environment fit theory to investigate social connections at work and mental health during the first wave of the COVID-19 pandemic. It analyses employees’ needs for social connections and how social connections affect different mental health measures. Survey data were collected in May 2020 in an online survey of employees across Germany and analysed using response surface analysis. Mental health was measured as positive mental health and mental health disorders. Social connections were measured as social support and social interactions. 507 employees participated in the survey and more than one third reported having less social support and social interaction at work than they desired (p < .001). This was associated with a decrease in mental health. In contrast, having more than the desired amount of social support was associated with a decrease and having more than the desired amount of social interaction with an increase in mental health. This study provides important early evidence on the impact of the first wave of the COVID-19 pandemic in the workplace. With it, we aim to stimulate further research in the field and provide early evidence on potential mental health consequences of social distancing measures – while also opening avenues to combat them.

## Introduction

The pandemic of coronavirus disease (COVID-19) has caused major problems for economies, health systems and societies around the world [1, 2]. In an attempt to slow the spread of the virus, governments have implemented quarantine, isolation and social distancing measures [3, 4]. These have limited social connections with friends, family and co-workers [3, 5] and changed the nature of social interactions [6]. Social connections, however, are a central part of human life, and both their quality and quantity can strongly influence mental health [7, 8]. Despite this, empirical evidence on the psychological, behavioural and social effects of the pandemic and the consequences for mental health, especially in the workplace, is sparse [9– 11]. This is striking given that the workplace is considered to be one of the most important environments to affect mental health [12] and recent studies suggest that the likelihood and duration of insufficient social relationships may be greater in the workplace than in personal life [7]. Research on social connections at work and mental health during all waves of COVID-19 is therefore urgently needed to better understand the consequences of the pandemic and develop appropriate countermeasures.

Some evidence on mental health consequences can be drawn from earlier epidemics, where quarantine was linked to an increase in the prevalence of depression, posttraumatic stress disorders and anxiety [13, 14]. Additionally, both quarantine and social distancing have been shown to be associated with various psychological stressors, such as frustration, boredom or inadequate information, which can have negative, sometimes long-lasting effects on mental health [5]. However, research on the mental health consequences of earlier epidemics and pandemics is sparse, and the extent and duration of the measures to contain COVID-19 are unprecedented [15]. In the context of the nationwide measures across most of the world, policymakers and researchers have raised concerns that people will experience prolonged social isolation [9, 16, 17], which can be defined as the absence or limitation of social connections [18]. Even though social isolation itself is associated with morbidity, related research suggests that mental health is less influenced by the objective absence of social connections than by the perceived discrepancy between actual and desired connections [16, 19]. When investigating the consequences of the COVID-19 pandemic in the workplace, it is therefore important to consider this discrepancy as a potential pathway through which COVID-19 and the corresponding containment measures could affect employees’ mental health.

One of the most common ways to examine perceived discrepancies is Person-Environment fit theory, in which fit is understood as the congruence, match or similarity between personal and environment factors [20]. Studies investigating the relationship between social connections and mental health suggest that the fit between a person’s needs for social connections and the supplies of the environment positively influence mental health. More specifically, mental health increases as the supplies of social interaction (i.e., in terms of quality and quantity) increase towards the needs of the person. When the supplies of the workplace fall short of a person’s needs, employees experience stress and loneliness, resulting in reduced mental health [21]. Excess social interactions, on the other hand, can interfere with the need for privacy or inhibit work that requires solitude, thereby reducing mental health [22, 23]. A similar effect can be found for social support. A greater mismatch between needed social support and supplied social support is associated with greater depressive symptoms but follows an asymmetric functional form where depressive symptoms are highest when needs exceed supplies [19].

Building on Person-Environment fit, our study investigates social connections at the workplace and employees’ mental health during the first wave of the COVID-19 pandemic. We analysed the extent to which employees experienced discrepancies in their social connections at work, and we investigated the relationship between these discrepancies and mental health during the pandemic. To capture the broad spectrum of mental health, we used measures for positive mental health and mental health disorders [24]. To capture social connectedness at work, we used measures of social support [25] and social interactions [26], which are two of the most important dimensions of social connectedness [26]. We employed polynomial regression and response surface analysis (RSA) to investigate the interplay between the two predictor variables (i.e., needs for and supplies of social connections) and their combined effect on mental health [27, 28].

The innovation of our study lies in its analysis of social connections in the workplace as a potential pathway through which COVID-19 and the corresponding containment measures may impact employees’ mental health. Understanding cause-effect relationships is essential to avoiding or mitigating unintended consequences through appropriate countermeasures. Our study provides policymakers and researchers around the world with important initial insights into social connections at the workplace and the potential consequences of social distancing interventions on employees’ mental health during the first wave of the COVID-19 pandemic. The conceptual model tested in this study is summarised in Figure 1.

**Fig 1.**
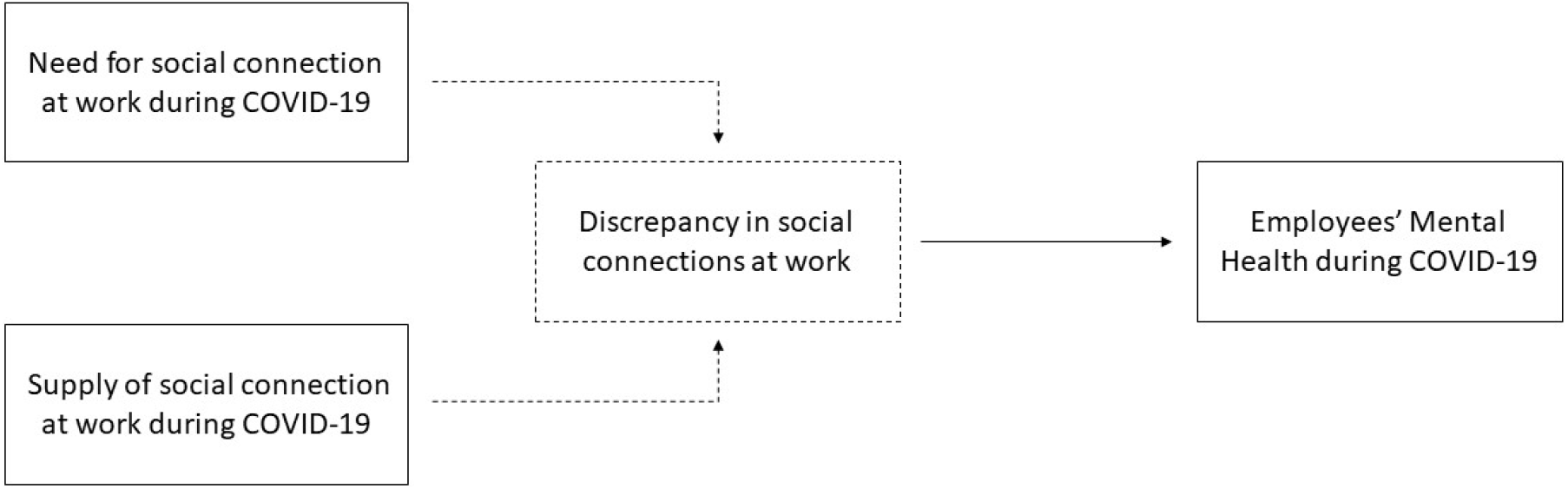
Conceptual Framework (Adapted from Bohndick and colleagues [29]).

## Methodology

### Study design and study population

Our research setting was the German workplace in the first wave of the COVID-19 pandemic, during which strict containment measures were in place and millions of employees had to work from home. A self-designed web survey was used to collect nationwide data on employees’ social connections at work (i.e., social interaction and social support) and mental health (i.e., positive mental health and mental health disorders). The data collection period was from 6 to 20 May 2020. Links to the survey were distributed through mailing lists and repeatedly posted on social media groups (mainly Facebook but also Xing, LinkedIn and Reddit) related to COVID-19. In total, 566 individuals completed the survey. Unreliably fast respondents (i.e., those who replied within fewer than two minutes, comprising 1.06% of the sample) [following 19] and participants who worked fewer than an average of five hours per week during the COVID-19 pandemic (9.36% of the sample) were excluded (N = 59), yielding a final sample of 507 participants.

### Measures

Validated and established scales from the literature were used to measure mental health and social connections at work [24, 30–32] (see ESM 1). The survey was conducted in German, and the widely accepted translation-back translation approach of Brislin [33] was used to ensure that the original items and scales retained their meaning.

#### Mental health

To capture the broad spectrum of mental health, we measured positive mental health and mental health disorders. To assess positive mental health, we used the Positive Mental Health Scale (PMH-9) of Lukat and colleagues [24]. This short, unidimensional scale consists of nine self-reported items to be rated on a four-point Likert scale ranging from “not applicable” (1) to “applicable” (4). Cronbach’s alpha for the PMH-9 was .91, indicating high internal consistency. To measure mental health disorders, we used the German version of the Patient Health Questionnaire (PHQ-4). The four-item PHQ-4 is a brief self-report questionnaire that assesses participants’ mental health using the last two weeks as a reference frame. It consists of a two-item depression (PHQ-2) and a two-item anxiety scale (GAD-2), both rated on a four-point Likert scale ranging from “never” (1) to “almost every day” (4). Studies have shown good reliability and validity of the PHQ-4 and its two subscales in the general population in Germany [32]. Cronbach’s alpha for the PHQ-4 for our study was .82.

#### Social connections at work

Social connections at work were captured with two dimensions highlighted by previous research: social support and social interaction [26]. For both dimensions, needs and supplies were assessed separately using commensurate scales [20]. Participants were asked to rate a) their needs for and b) their supplies of social connections at work on five-point Likert scales ranging from “very rarely” (1) to “very often” (5). Items from Ehrhardt and Ragins [31] needs-supplies scale for instrumental and personal support were used. The scale contains four items for needs and supplies and has shown good validity in previous studies [31]. Cronbach’s alpha for both needs and supplies in our study was .86. To measure social interaction at work, we used the widely employed and accepted [34, 35] social need non-fulfilment subscale of Cook and Wall [30]. This four-item scale is adapted to assess the items on commensurate scales. This approach of constructing commensurate measures based on well-established scales has been used in numerous studies of P-E fit [23]. Cronbach’s alpha was .95 for needs and .93 for supplies in our study.

#### Control variables

To control for demographic and work-related characteristics that might affect our outcome variables but were not pertinent to our primary research question, we included the following control variables: gender, age, marital status, years of education, occupation, and weekly work-hours during the first wave of COVID-19.

### Analytical strategy

First, descriptive statistics including mean, standard deviation and frequency distributions were computed. Paired samples t-tests and algebraic difference scores were used to analyse differences between needs and supplies for social connections (i.e., social support and social interactions).

Second, the relationship between social connections at work and mental health was analysed using Response Surface Analysis (RSA) (see also ESM 2). RSA enables researchers to test complex psychological effects and investigate whether the congruence of two constructs (in this case, needs for and supplies of social connections) is associated with higher values in mental health [36]. RSA is the state-of-the-art methodology for investigating these relationships and can identify more elaborate effects than comparable methods (e.g., difference scores) [36]. For this, the R package RSA version 0.9.13 was used [37]. Predictor variables for needs and supplies were standardised by centring them at their grand mean and then dividing them by their grand standard deviation [36]. Multicollinearity between the two predictors was adequately low, with a variance inflation factor below two. No multivariate outliers according to the Bollen and Jackman [38] criterion were detected. For the RSA, an unconstrained polynomial regression model of second degree was calculated and visualised in a three-dimensional surface plot. Fit effects were investigated using the checklist by Humberg and colleagues [36]. Following earlier research [36, 39], the regression models and surface plots were used to investigate the meaning and magnitude of the effects more closely.

A declaration of compliance with terms of use and ethical standards from the University of Hamburg’s WiSo Laboratories was obtained on 03 March 2022 (no approval number provided). Upon opening the questionnaire, participants were informed that their participation was voluntary and that their anonymised data would be used for scientific purposes only. Informed consent was not required because the questionnaires were answered anonymously. All calculations were performed using R, version 3.6.1 [40]. The significance level was set at *p =* .*05*.

## Results

### Demographic characteristics

Participants had a mean value of 1.84 (*SD=0*.*64*) for mental health disorders and 2.88 (*SD=0*.*66*) for positive mental health. More demographic and work-related characteristics of the participants are presented in Table 1.

**Table 1.** Demographic characteristics of the participants

| Variables | Mean | SD | Minimum | Maximum |
| --- | --- | --- | --- | --- |
| Age (in years) | 38.49 | 12.03 | 18 | 80 |
| Gender (in %) |  |  |  |  |
| Male | 31.76 |  |  |  |
| Female | 68.05 |  |  |  |
| Other | 0.20 |  |  |  |
| Education (in years) | 16.04 | 4.50 | 0 | 34 |
| Marital Status (in %) |  |  |  |  |
| Single | 49.51 |  |  |  |
| Married | 41.82 |  |  |  |
| Divorced | 6.11 |  |  |  |
| Widowed | 1.18 |  |  |  |
| Weekly work hours | 32.38 | 11.92 | 5 | 70 |
| Mental health disorders (PHQ-4) | 1.84 | 0.64 | 1 | 4 |
| Positive mental health (PMH-9) | 2.88 | 0.66 | 1 | 4 |

*Note.* $N = 507$ ; $SD$ = Standard Deviation

### Discrepancies in social connections during COVID-19

Paired samples t-tests revealed that the needs for social interaction at work were significantly higher than its supplies (*t* = 10.10, *p* < .05). Additionally, the needs for social support at work were significantly higher than its supplies (*t* = 8.54, *p* < .05). This suggests that, on average, individuals’ needs for social support and social interaction exceeded the amount supplied at the workplace, indicating a mismatch in the general population of the sample.

To further analyse this mismatch, difference scores were created. Following Rankin and colleagues [19] and Fleenor and colleagues [41], difference scores greater than half a standard deviation were considered discrepant. During the first wave of the COVID-19 pandemic, 43.98% (*n* = 223) of participants had fitting needs-supplies parameters for social interaction, and 32.54% had fitting needs-supplies parameters for social support (*n*=165). Furthermore, 41.81% (*n* = 212) of participants reported that they had less social interaction at work than they desired, and 37.48% (*n* = 190) of participants reported that they received less social support at work than they desired.

### Response Surface Analyses

RSA was conducted to determine the relationship between social connections at work and mental health during the first wave of the COVID-19 pandemic. Four polynomial regression models were calculated and visualised in three-dimensional surface plots. The explained variance of the models ranged from 9.6% (model 4) to 10.9% (model 1), which is in line with similar studies using RSA [23, 27]. The standardised polynomial regression coefficients are shown in Table 2, and the surface parameters are presented in Table 3. The surface plots are depicted in Figure 2.

**Table 2.** Estimated polynomial regression model

| | $b_0$ | $b_1$ | $b_2$ | $b_3$ | $b_4$ | $b_5$ |
| --- | --- | --- | --- | --- | --- | --- |
| a) Social interaction and positive mental health | 2.98*<br>[2.90; 3.06] | 0.18*<br>[0.10; 0.25] | -0.06<br>[-0.14; 0.01] | -0.01<br>[-0.06; 0.05] | 0.06<br>[-0.02; 0.14] | -0.04<br>[-0.12; 0.03] |
| b) Social interaction and mental health disorders | 1.74*<br>[1.66; 1.82] | -0.19*<br>[-0.25; -0.13] | 0.08*<br>[0.01; 0.14] | 0.00<br>[-0.05; 0.05] | -0.02<br>[-0.08; 0.05] | 0.04<br>[-0.02; 0.10] |
| c) Social support and positive mental health | 2.95*<br>[2.87; 3.03] | 0.16*<br>[0.09; 0.22] | -0.14*<br>[-0.21; -0.07] | -0.06<br>[-0.12; 0.00] | 0.08*<br>[0.00; 0.15] | 0.02<br>[-0.05; 0.08] |
| d) Social support and mental health disorders | 1.78*<br>[1.70; 1.86] | -0.14*<br>[-0.21; -0.07] | 0.12*<br>[0.05; 0.19] | 0.36<br>[-0.03; 0.10] | -0.09*<br>[-0.16; -0.02] | 0.01<br>[-0.05; 0.07] |

*Note.* Full polynomial regression model $Z = b_0 + b_1 * X + b_2 * Y + b_3 * X^2 + b_4 * XY + b_5 * Y^2$ ; $X$ = supplies; $Y$ = needs; Columns show regression coefficient estimates and confidence intervals [in square brackets]; \* $p < .05$

**Table 3.** Response surface results

|  | First principal axis |  | Line of congruence |  | Line of incongruence |  | Conclusion |
| --- | --- | --- | --- | --- | --- | --- | --- |
| | $p_{10}/p_{20}$ | $p_{11}/p_{21}$ | $a_1$ | $a_2$ | $a_3$ | $a_4$ | |
| a) Social interaction and positive mental health | -1.30<br>[-4.18; 1.37] | 0.58<br>[-0.19; 1.35] | 0.11*<br>[0.03; 0.19] | 0.01<br>[-0.06; 0.08] | 0.24*<br>[0.11; 0.37] | -0.11<br>[-0.26; 0.04] | No congruence<br>($a_4=0$, $a_3 \neq 0$) |
| b) Social interaction and mental health disorders | -1.43<br>[-4.08; 1.23] | 0.21<br>[-0.58; 1.00] | -0.11*<br>[-0.19; -0.04] | 0.02<br>[-0.05; 0.09] | -0.27*<br>[-0.37; -0.17] | 0.06<br>[-0.07; 0.189] | No congruence<br>($a_4=0$, $a_3 \neq 0$) |
| c) Social support and positive mental health | -3.55<br>[-8.93; 1.83] | 2.44<br>[-0.44; 5.32] | 0.02<br>[-0.05; 0.08] | 0.04<br>[-0.03; 0.10] | 0.29*<br>[0.17; 0.41] | -0.11<br>[-0.25; 0.03] | No congruence<br>($a_4=0$, $a_3 \neq 0$) |
| d) Social support and mental health disorders | -2.13<br>[-5.00; 0.73] | 1.34<br>[-0.06; 2.75] | -0.02<br>[-0.08; 0.05] | -0.05<br>[-0.11; 0.02] | -0.26*<br>[-0.38; -0.13] | 0.14*<br>[0.00; 0.27] | No congruence<br>($a_3 \neq 0$) |

*Note.* Position of the first principal axis = $p_{10} + p_{11}X$ ; LOC = Line of congruence; Shape of the surface above the LOC $Z = b_0 + a_1X + a_2X^2$ ; LOIC = Line of incongruence; Shape of the surface above the LOIC $Z = b_0 + a_3X + a_4X^2$ ; Columns show regression coefficient estimates and confidence intervals [in square brackets]; \* $p < .05$

**Fig 2.**
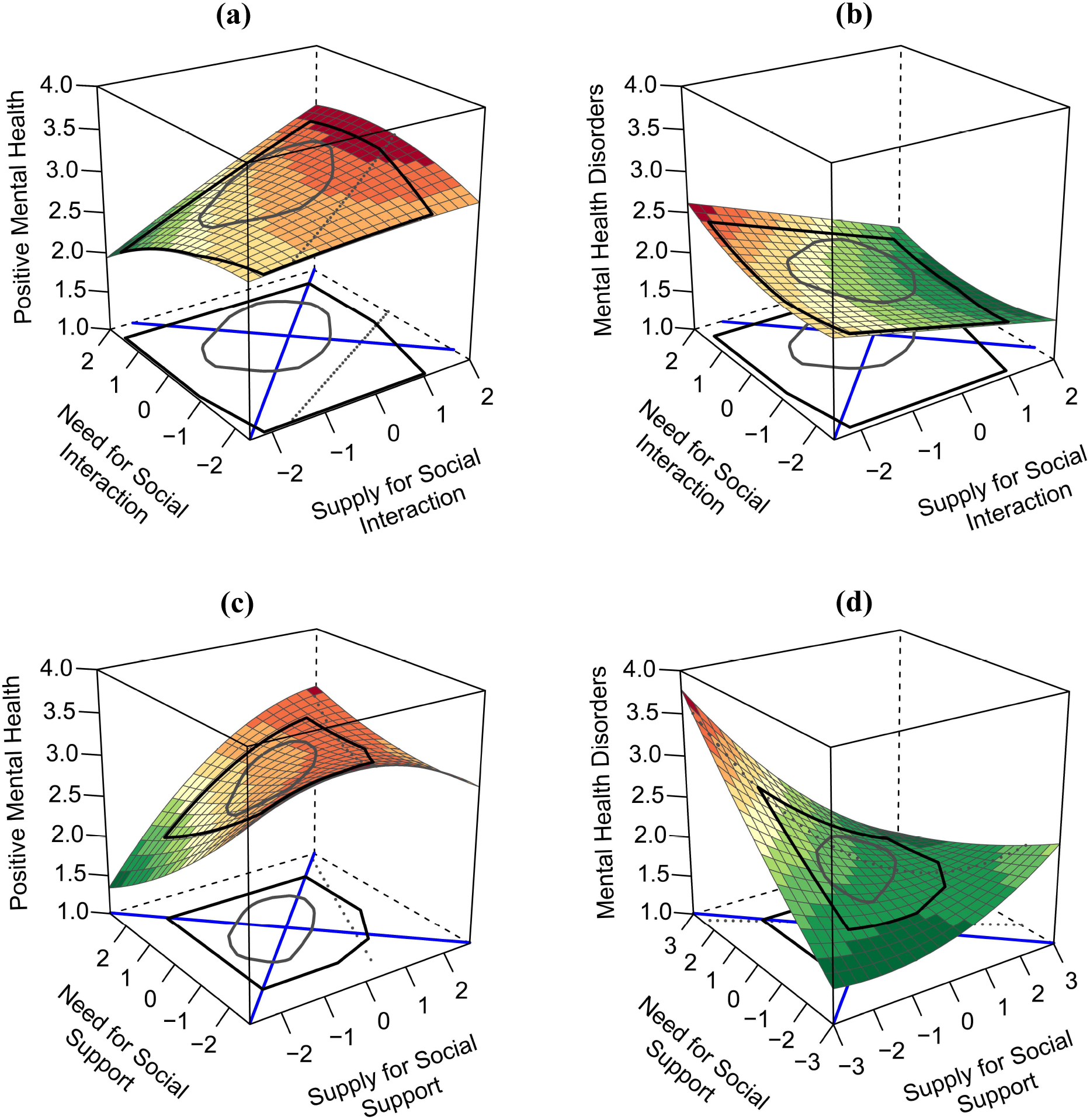
The impact of social connection measures on measures of mental health, displayed in four response surface plots. *Note*. Surface plots a) and b) assess the needed social interaction (*x* axis) and supplied social interaction (*y* axis), and c) and d) assess the needed social support (*x* axis) and supplied social support (*y* axis). Surface plots a) and c) predict positive mental health (*z* axis), and b) and d) predict mental health disorders (*z* axis). Colour scheme: Darker shades of red indicate higher values within each plot, and darker green shades indicate lower values.

First, all four models were evaluated for fit effects using the checklist by Humberg and colleagues [36]. In all cases, at least one condition was violated and the RSA contradicted a fit effect. This finding indicates that people whose needs fit their supplies were not the ones with the highest positive mental health (models a & b) or the lowest level of mental health disorders (models c & d). Even though no fit effect could be found^1^, our analyses of the surface plot and regression coefficients allowed us to interpret the meaning and magnitude of the effects more closely [36].

Linear main effects of the needs (for models b, c and d) and the supplies (for all models) seemed to be the most prominent and consistent patterns in our data (see Table 2). The supplies (*b*_1_) of social interaction and social support had a positive influence on positive mental health and a negative influence on mental health disorders. The needs (*b*_2_) for social interaction had no significant effect on positive mental health and a positive effect on mental health disorders, whereas the needs for social support had a negative effect on positive mental health and a positive effect on mental health disorders. For all models, the effect size of the supplies was stronger than the effect size of the needs. This finding was especially pronounced for models a) and b), resulting in a tilted surface for both plots. Additionally, the higher-order terms (*b*_3_ - *b*_5_) in both models had no significant influence on the outcome variables, resulting in plane surface plots.

In contrast, the interaction term (*b*_4_) showed a significant influence in models c) and d), indicating a joint influence of needs for and supplies of social support on mental health. People who rated their needs for social support as high and their supplies as low (left corner in Figure 1d) reported the highest level of mental health disorders. In contrast, people who rated their supplies of social support as high and their needs as low (right corner) reported a moderate level of mental health disorders, whereas people with fitting parameters (on the ridge of the surface) reported a low level of mental health disorders. Due to the positive a_4_ coefficient, needs for social support at work that exceeded supplies led to a higher level of mental health disorders than supplies that exceeded needs. For model c), people who rated both needs and supplies as high (back of the cube) reported the highest level of positive mental health. In contrast, people with moderate needs and supplies (centre of the cube) whose supplies exceeded their needs reported the highest level of positive mental health.

### Sensitivity analyses

To explore the robustness of the results, two sensitivity analyses were conducted (see ESM 3). First, all four polynomial regression models were re-estimated using the full sample of 566 participants. Second, models b) and d) were re-estimated using the PHQ-2 instead of the PHQ-4 to measure mental health disorders. Across both sensitivity analyses, effects did not change in sign, size or significance, indicating that our results are robust.

## Discussion

Building on Person-Environment fit theory, we investigated the relationship between social connections at work and employees’ mental health during the first wave of the COVID-19 pandemic. Our results indicate that employees’ needs for social connections in the workplace were significantly higher than the supplies of these connections. Indeed, more than one third of our sample received less social support and social interaction at work than they desired. This study is one of the first to measure employees’ needs for and supplies of social connections during COVID-19 and to call attention to a significant lack of social interaction and social support in the workplace. This result is in concordance with other studies that have found, more generally, high levels of social isolation and loneliness [3, 42], as well as changes in social life and social connections [4, 43, 44], during the pandemic.

Furthermore, our response surface analysis (RSA) challenges the assumption of a fit effect, which is common in previous studies [22]. Instead, we found notable main effects in the first set of RSA models. Both models suggest a linear relationship between social connections at work and mental health, with mental health improving as supplies increase towards the level of needs and continuing to improve as they exceed it. However, social interactions during the first wave of the COVID-19 pandemic differ substantially from those under general conditions. Our results might indicate that this change in conditions has altered the relationship between social connections at work and mental health. Most notably, there appears to be no point during the pandemic at which social interactions at work become “too much” and reduce mental health. Prior research (e.g., French and colleagues [45], who investigated 2010 employees of 23 different occupations) suggests that an excess of social interaction can reduce mental health by impairing the need for privacy or inhibiting work that requires solitude. One possible explanation for our results might be found in the theory of carry-over effects. As suggested in prior work by Edwards and Rothbard [22], excess supplies can be conserved as a social resource and used to fulfil a lack of these supplies in other content dimensions. It seems plausible that during the first wave of the COVID-19 pandemic, an excess of social interactions at work might have been used to fulfil unmet needs for social interaction in other areas of life (e.g., at home).

Furthermore, the results of our study suggest that the level of mental health disorders are lowest when employees’ social support at work matches their preferences. In our sample, when supplies started to exceed needs, however, the level of mental health disorders increased gradually. A similar but opposite effect could be found for positive mental health. These findings suggest that when social support exceeds an employee’s needs, the positive effects of the additional support and any carry-over effects are overshadowed by the intrusion this represents into the employee’s need for privacy [22]. Even though no fit effect could be found in this regard, these results are supported by the literature [19, 22]. The results of our study add evidence in support of this hypothesis and extend the existing literature by showing a similar effect on positive mental health. Our results indicate that a level of social support at work that fits the needs of the employee might be an important resource to improve mental health during the first wave of the COVID-19 pandemic.

Although our study makes a number of important contributions, our results should be considered in light of certain limitations, each of which provides an avenue for future research. First, our study investigates only the impact of the first wave of the COVID-19 pandemic in the workplace. At the time of our study, containment measures and changes to work environments, such as remote working and video conferencing, were still new and unfamiliar to many employees. During later waves of the pandemic, however, employees may have become accustomed to these new conditions and changed their perceptions of social support and social interaction at the workplace. As a result, findings from different time points during the pandemic might vary due to habituation effects, as well as changes in the length and severity of restrictions. Further research of a longitudinal nature, ideally with various data points before, during and after the pandemic would allow more reliable statements to be made on causality, habituation, and changes in social connections at work and mental health during the pandemic. Second, the cross-sectional design does not allow conclusions to be drawn about causality. A third limitation concerns our setting and sample. Our data are from an online survey, which allowed us to recruit a large number of participants efficiently in a short period. However, online surveys are likely to miss participants that are vulnerable to the pandemic, including people with no internet access or in rural communities [46]. Future research is needed to better include these groups. Although growing evidence suggests that social media is a useful tool for recruiting participants for health research (e.g., because it reduces costs and recruitment time) [47], it can introduce self-selection biases [29]. Despite the fact that our sample was relatively large (*n=507*), it is not representative and differs from the general working population in Germany, especially in terms of its small proportion of male respondents (31.76% male) and large proportion of highly educated people (mean of 16.04 years of education) [48]. These aspects might be a threat to external validity and restrict the generalisability of our results. Given the importance of this topic, further research replicating our analyses in different settings (e.g., cross-country evidence) with large, representative samples is needed.

## Supporting information

Electronic Supplementary Material

## Data Availability

All files are available on OSF.io.

## Conclusion and implications

Building on Person-Environment fit theory, our study provides new insights into social connections at work and mental health during the first wave of the COVID-19 pandemic. The needs for social connections in our sample of employees were significantly higher than the supplies of these connections in the workplace, and more than every third employee received less social interaction and social support than they desired. This could have important mental health implications given that employees whose supplies of social connections were below their needs reported low levels of mental health. As supplies increase towards the needs, employees’ mental health increased. Even excess social interactions were associated with an increase in mental health, possible because they could help fulfil employees’ needs for social interaction in other areas of life. Too much social support, however, was associated with a decrease in mental health, potentially because it impinged upon employees’ needs for privacy.

These findings have significant **practical implications** as they provide initial evidence on two avenues that could help combat the potential mental health consequences of social distancing measures. Instead of providing an arbitrary amount of social support, businesses, policymakers and managers might seek to create a work environment in which their employees receive a level of social support that aligns more closely with the amount that the employees feel they need. Too little social support at work can result in loneliness, whereas too much can interfere with the needs for privacy – both negatively affecting mental health. Second, an increase in social interactions at work can lead to better mental health. Our findings call attention to the idea that social interactions at work that are in excess of what employees indicate as their needs might carry their positive effect over into other areas of life, improving employees’ mental health during the first wave of the pandemic. This provides important information for researchers and policymakers in the field of mental health. In particular, a continuous needs-supplies assessment through instruments such as standardized employee surveys could help reveal potential discrepancies and identify where measures might be taken to improve employees’ social connections and mental health.

## Footnotes

1 Even if no fit effect can be found, the results of the surface plot can still be interpreted. Beyond fit effects, RSA can address a wide range of further questions – for example, additional main effects or curvilinear main effects [36].

