## Supplementary material for "Social connections at work and mental health during the first wave of the COVID-19 pandemic: Evidence from employees in Germany": Electronic Supplementary Material

### ESM 1: Measurement instruments

Measurement instruments including all items used in this study can be found in Table S1. The internal consistency of all constructs was good, with Cronbach’s alpha values ranging between α = .82 and α = .95.

| Construct and Items | | |  | | |
| --- | --- | --- | --- | --- | --- |
| *Positive Mental Health (9 Items)*  I am often carefree and in good spirits; I enjoy my life; All in all, I am satisfied with my life; In general, I am confident; I manage well to fulfil my needs; I am in good physical and emotional condition; I feel that I am actually well equipped to deal with my life and its difficulties; Much of what I do brings me joy; I am a calm, balanced human being. | | | *α* = .91  *M* = 2.88  *SD* = 0.66 | | |
| *Mental health disorder (4 Items)*  Little interest or pleasure in doing things; Feeling down, depressed or hopeless; Feeling nervous, anxious or on edge; Not being able to stop or control worrying. | | | *α* = .82  *M* = 1.84  *SD* = 0.64 | | |
|  | | | Need | Supply | |
| *Social relationships at work (4 Items)*  The opportunity to talk to others; The opportunity to make friends; being part of a social group; friendly contact with others. | | | *α* = .86  *M* = 3.69  *SD* = 0.89 | | *α* = .86  *M* = 3.15  *SD*=1.08 |
| *Social support at work (4 Items)*  provide you with support on personal matters; offer you help on personal issues or challenges; offer to listen to a problem you may be having; go out of their way to help you with personal issues. | | | *α* = .95  *M* = 3.25  *SD* = 1.05 | *α* = .92  *M* = 2.81  *SD* =1.13 | |

**Table S1** Measurement Instruments

*Note.* *α* = Cronbach’s alpha, *M* = Mean, *SD* = Standard Deviation

### ESM 2: Respon**se Surface Analysis**

Since the application of RSA is still rare (Milatz et al., 2015), a short overview of the RSA methodology and our analytical process is given below. The RSA consists of two steps (Humberg et al., 2019). In the first step, the following unconstrained polynomial regression model of second-degree, including the two predictor variables (*X* and *Y*), their squared terms *(*$X^{2}$ and $Y^{2}$), and the interaction of both predictors (*X*Y*) is calculated (Milatz et al., 2015; Nestler et al., 2019):

$$Z=b_{0}+b_{1}X+b_{2}Y+{b_{3}X^{2}+b_{4}Y^{2}+b}_{5}XY+\varepsilon$$

In the second step of the RSA, a three-dimensional graph of the regression model is created. The graph in combination with the regression coefficients and surface parameters is used to interpret the estimated polynomial regression model and to investigate the meaning of the effects more closely. (Humberg et al., 2019; Milatz et al., 2015).

To further guide the interpretation, the checklist of Humberg et al. (2019) was used to identify fit effects (also called congruence effects). To detect fit effects, three features of the response surface are especially important: The ridge of the surface (called First Principal Axis; FPA), the line on the XY plane that contains all congruent predictor combinations *Y = X* (called Line of Congruence; LOC), and the line on the XY plane that contains all incongruent predictor combinations *Y = -X* (called Line of Incongruence; LOIC) (Humberg et al., 2019; Nestler et al., 2019). These features must be considered jointly (Nestler et al., 2019) and have to satisfy at least four conditions to reflect a fit effect (Humberg et al., 2019). To represent a fit effect, the response surface needs to predict the highest outcome for people with congruent predictors (Schönbrodt, 2016). For this, the FPA must not differ significantly from the LOC (Humberg et al., 2019). This is the case when the intercept of the FPA ($p_{10}$) is not significantly different from 0 (condition 1) and the slope of the FPA ($p_{11}$) is not significantly different from 1 (i.e. the confidence interval of $p_{11}$ should include 1; condition 2) (Humberg et al., 2019). Additionally, people with increasingly incongruent predictors need to have significantly lower outcome values (Schönbrodt, 2016). This is the case when the slope of the LOIC at the origin ($a_{3}$) does not significantly differ from 0 (condition 3) and the quadratic term coefficient ($a_{4}$) is significantly negative (condition 4) (Humberg et al., 2019). In sum, the four conditions for fit effects are: (1) $p_{10}$ $\approx$0, (2) $p_{11} \approx$1, (3)$a_{4}$ $<$ 0, and (4) $a_{3}$ $\approx$0 (Humberg et al., 2019). If one of the four conditions is violated, the fit hypothesis must be rejected (Humberg et al., 2019).

### ESM 3: Sensitivity analysis

Results of the sensitivity analysis can be found in Table S2 and Table S3.

**Table S2** Sensitivity analysis one: Re-estimated polynomial regression model with the full sample

|  | $b_{0}$ | $b_{1}$ | $b_{2}$ | $b_{3}$ | $b_{4}$ | $b_{5}$ |
| --- | --- | --- | --- | --- | --- | --- |
| a) Social interaction and positive mental health | 3.00*  [+0.02] | 0.20*  [+0.02] | -0.05  [-0.01] | -0.03  [+0.02] | 0.02  [+0.02] | -0.06  [+0.02] |
| b) Social interaction and mental health disorders | 1.73*  [-0.01] | -0.19*  [0.00] | 0.08*  [0.00] | 0.02  [+0.02] | 0.00  [-0.02] | 0.05  [+0.01] |
| c) Social support and positive mental health | 2.96*  [+0.01] | 0.18*  [+0.02] | -0.12*  [+0.02] | **-0.07***  [-0.01] | 0.07*  [-0.01] | 0.01  [-0.01] |
| d) Social support and mental health disorders | 1.77*  [-0.01] | -0.15*  [-0.1] | 0.10*  [-0.02] | 0.05  [-0.31] | -0.08*  [+0.01] | 0.03  [+0.02] |

*Note.* Full polynomial regression model $Z=b_{0}+b_{1}*X+b_{2}*Y+b_{3}*X^{2}+b_{4}*XY+b_{5}*Y^{2}$; *X* = supplies; *Y* = needs; Columns show regression coefficient estimates and coefficient changes in comparison to the original model [in parenthesis]; significance changes are indicated in **bold**; * *p* < .05

**Table S3** Sensitivity analysis two: Re-estimated polynomial regression model using the PHQ-2 to assess mental health disorders

|  | $b_{0}$ | $b_{1}$ | $b_{2}$ | $b_{3}$ | $b_{4}$ | $b_{5}$ |
| --- | --- | --- | --- | --- | --- | --- |
| b) Social interaction and mental health disorders | 1.82*  [+0.08] | -0.21*  [-0.02] | 0.10*  [+0.02] | 0.00  [0.00] | -0.02  [0.00] | 0.03  [-0.01] |
| d) Social support and mental health disorders | 1.86*  [+0.08] | -0.15*  [-0.01] | 0.14*  [+0.02] | 0.36  [-0.00] | -0.10*  [-0.01] | 0.01  [0.00] |

*Note.* Full polynomial regression model $Z=b_{0}+b_{1}*X+b_{2}*Y+b_{3}*X^{2}+b_{4}*XY+b_{5}*Y^{2}$; *X* = supplies; *Y* = needs; Columns show regression coefficient estimates and coefficient changes in comparison to the original model [in parenthesis]; significance changes are indicated in **bold**; * *p* < .05
